# SARS-CoV-2 testing of aircraft wastewater shows that mandatory tests and vaccination pass before boarding did not prevent massive importation of Omicron variant in Europe

**DOI:** 10.1101/2022.04.19.22274028

**Authors:** Lorlane Le Targa, Nathalie Wurtz, Alexandre Lacoste, Gwilherm Penant, Priscilla Jardot, Alexandre Annessi, Philippe Colson, Bernard La Scola, Sarah Aherfi

**Affiliations:** MEPHI, Institut de Recherche pour le Développement (IRD), Assistance Publique - Hôpitaux de Marseille (AP-HM), Aix-Marseille Université, Marseille, France; IHU Méditerranée Infection, Marseille, France; Biosellal; Bataillon des Marins Pompiers de la ville de Marseille, Marseille, France

**Author notes:** **Corresponding authors:** Sarah Aherfi, IHU - Méditerranée Infection, 19-21 boulevard Jean Moulin, 13005 Marseille, France., Bernard La Scola, IHU - Méditerranée Infection, AP-HM, 19-21 boulevard Jean Moulin, 13005 Marseille, France.

**Keywords:** SARS-CoV2, wastewater monitoring, variants spreading, aircraft, surveillance

## Abstract

**Background:** Most new SARS-CoV-2 epidemics in France occurred following importation from abroad of emerging viral variants. Currently, the control of such risk of new variant importation is based on the negativity of a screening test (PCR or antigenic) and on an up-to-date vaccine status, such as International Air Transport Association travel pass.

**Methods:** Wastewater of 2 planes arriving in Marseille (France) from Addis-Ababa (Ethiopia) on December 2021 were i) tested by RT-PCR for SARS-CoV2 detection, and variants screening; these tests were carried out between landing and custom clearance, ii)sequenced by MiSeq Illumina.

Antigenic tests and sequencing by NovaSeq were carried out on respiratory samples collected from the 56 passengers of the second flight.

**Results:** SARS-CoV-2 RNA suspected of being from the Omicron BA.1 variant was detected on the aircraft’s wastewater.,

SARS-CoV2 RNA was detected for 11 (20%) passengers and the Omicron BA.1 variant was identified.

**Conclusion:** Our work shows the efficiency of aircraft wastewater testing to detect SARS-CoV-2 cases among travelers and identify the viral genotype. It also highlights the low performance for incoming flights from outside Europe to France of the current filter strategy that combines requirement for a vaccine pass and a negative testing before boarding.

## Introduction

Coronavirus disease-2019 (COVID-19), caused by the Severe Acute Respiratory Syndrome – Coronavirus 2 (SARS-COV-2), emerged in Wuhan (China) in December 2019 and since then became a pandemic with more than 336□million confirmed cases globally and 5.5 million deaths as of January 20, 2022. Most SARS-CoV-2 epidemics that occurred successively or concurrently at the level of countries resulted from the importation from abroad of emerging viral variants [1]. Indeed, air travel and boat cruise have been associated with the spread of SARS-CoV-2 including of new variants via infected passengers. Since the beginning of the COVID-19 pandemic, many countries and regions imposed restrictions, for example quarantines, entry bans, obligation of vaccination, or travel restrictions. To know the restrictions imposed in each country, the International Air Transport Association (IATA) developed a COVID-19 Travel Regulations Map (powered by Timatic), which gives in real time the requirements according to the itinerary with certainty for anywhere in the world (https://www.iatatravelcentre.com/world.php). Previous studies have examined the effect of travel restrictions and travel-related measures imposed during the pandemic. Most of these studies focused on the initial phase of the COVID-19 spread, when the epidemic was concentrated in Wuhan (China) [2;3]. All of these studies found that travel restrictions in the early part of the epidemic helped to delay the spread of COVID-19. Other studies found that the restrictions were insufficient to control the global spread completely [4;5]. With the emergence of new SARS-CoV-2 variants, many countries have strengthened border control measures, with pre-travel and post-travel screening tests, to avoid their importation. In the last months, new variants had spread worldwide including most recently the Omicron variant that was first described in South Africa and Bostwana [6]. Its clinical manifestations are similar to those of other respiratory viral infections with dry cough, fever, tiredness, myalgia and difficulty to breath [6], but can also include gastrointestinal symptoms such as diarrhea, nausea, abdominal pain and vomiting in 2-10 % of cases [7]. High concentrations of SARS-CoV-2 RNA have been found in stools of infected asymptomatic and symptomatic people [8] and viruses were still infectious [9]. Therefore, analysis of SARS-CoV-2 in wastewater appears an interesting approach to monitor the disease burden in communities. After the first report of the detection of SARS-CoV-2 in wastewater by Medema et al. in Netherlands [10], detection and monitoring in wastewater samples have been reported in many countries [11-14]. A few studies have performed SARS-CoV-2 genome sequencing from sewage to identify viral genotypes circulating in a community and study genetic diversity [14-19]. Some of them showed congruence bewteen variants found in clinical isolates during the same period, while others identified genotypes not yet reported in clinical samples.

Until now, one study showed early in the pandemic that SARS-CoV-2 RNA was detected in wastewater from passenger aircraft [20;21] and this surveillance demonstrated a high positive predictive value for SARS-CoV-2 infection among people [22]. Only one study recently reported the successful detection by genome sequencing of variants in aircraft wastewater [23]. In the current study, we report the detection of SARS-CoV-2 variants in wastewater of aircrafts travelling from Addis Abeba, Ethiopia, to France. Two methods including full-length genome sequencing and real time reverse transcription (RT)-PCR (qPCR) to detect different variants by using the Bio-T Kit® FiveStar Covid-19 (Biosellal, Dardilly, France) were used. The high concentration of the Omicron variant in aircraft wastewater that was confirmed by antigenic testing of travelers shows massive importation of the Omicron variant in France from Africa. This confirms that the surveillance of aircraft wastewater provides precious public health information on the global spread of emerging SARS-CoV-2 variants and shows that production of a negative SARS-CoV-2 detection test before boarding does not guarantee that passengers are not viral carriers.

## MATERIALS AND METHODS

### Samples

A volume of 100 mL of aircraft wastewater samples from 2 flights arriving from Addis Abeba (Ethiopia) to Marseille (France) on December 22^nd^ and 24^th^ (indentified as 2212 and 2412 respectively) were collected on the aircraft of the airport tarmac by the bataillon des marins pompiers de Marseille (BMPM), and then stored at 5°C until the arrival at the laboratory. Samples were first passed through a paper filter to remove large particles, then a volume of 30 mL of the filtrate was filtered on a Millex sterile syringe filter with a pore size of 5 µm (SLSV025LS, Merck Millipore, MA, USA).

For the flight of December 24^th^, 2021, all passengers were proposed to have a nasopharyngeal swab sampled for SARS-CoV-2 detection according to a joint initiative of the regional prefecture and the regional health agency and according to decree No. 2020-551 of 12 May 2020 on the information systems mentioned in Article 11 of Law No. 2020-546 of 11 May 2020 extending the state of health emergency for people arriving from countries classified at risk if the virus circulates actively [24]. These nasopharyngeal swabs were sampled by the staff of BMPM for 56 passengers and tested by a rapid antigenic diagnosis test COVID-VIRO®, AAZ (Boulogne-Billancourt, France). For all patients tested positive, samples were supplied to our laboratory at +4°C for further RT-PCR and sequencing.

### Nucleic acids extraction

For wasterwater sample, before DNA/RNA extraction, 10µL of Bio-T Kit® FiveStar Covid-19 internal positive control (Biosellal, Dardilly, France) and 10 µL of magnetic silica are added in each sample. Nucleic acids from 1mL of each wastewater sample were extracted with the eGENE-UP^®^ Lysis and RNA/DNA Purification (Biomérieux, Marcy l’Etoile, France) to obtain a volume of 100µL of eluate. Negative control consisted in RNase Free water extracted following the same protocol.

For clinical samples, viral RNAs were extracted using the KingFisher Flex system (Thermo Fisher Scientific, Waltham, MA, USA) following the manufacturer’s recommendations.

### RT-PCR detection and variants screening

Direct Screening of SARS-CoV-2 variants in wastewater was performed on a RT-PCR Quantstudio5 device (Thermofisher, France) by using combination of the Bio-T Kit FiveStar Covid-19 and the Bio-T kit “Environmental Δ & O” (Biosellal, Dardilly, France). Confirmation of SARS-CoV-2 positivity attested by antigenic test in patients was confirmed by RT-PCR as previously described [25].

### Samples preparation for NGS sequencing

The first RNA/DNA extract from the sample 2212 was used without pre-treatment for further RT-PCR. For the sample 2412, 1 mL was freeze-dried then rehydrated in 30 µL of water. The reverse transcription step was performed in duplicate with the SuperScript VILO cDNA synthesis kit (11754-250, Thermo Fisher, Waltham, MA, USA) according to the supplier’s recommendations in a final volume of 20 µL per reaction. The ARTIC v3 PCR (ARTIC nCoV-2019 V3 Panel and 500rxn of IDT 10006788, Integrated DNA Technologies, Inc., Coralville, IA, USA) was carried out under the following conditions for one reaction for each of the pools 1 and 2: 2.5 µL of reaction Mix 10X, 0.5 µL dNTP (10 mM), 0.4 µL of forward primer (100 nM), 0.125 µL of HotStart qDNA Polymerase (Qiagen 203205, Hilden, Germany), water PCR grade (qsp 25µL) and 2 µL of template. Eight replicates were made per extract and per pool. The 8 replicates were then pooled (final volume of 200 μL) before purification on a Nucleofast 96 plates (Macherey Nagel ref 743100.50, Hoerdt, France). The purification products were eluted in 30 μL of TE 1X then placed on a 2% agarose gel (migration for 30 minutes, 100 V). The 400 base pair (bp) bands were cut out of the gel and purified according to the supplier protocol with the Monarch DNA Gel Extraction Kit (New England BioLabs, ref T1020L, Evry-Courcouronnes, France) with a final elution volume of 40µL.

For passengers’ samples, cDNA were amplified using with the Illumina COVIDSeq protocol including a multiplex PCR protocol with ARTIC nCoV-2019□V3 Panel primers (Integrated DNA technologies) according to the ARTIC procedure (https://artic.network/).

### NGS sequencing

For wastewater samples, final purification products were sequenced with the paired-end strategy with the Nextera XT DNA sample preparation kit (Illumina Inc, San Diego, CA, USA). These samples were barcoded for mixing with other projects. Libraries were prepared following the Illumina protocol (Illumina Inc, California, USA). PCR amplification to complete tag adapters and introduce dual index barcodes was done over 12 cycles followed by purification with 0.8x AMPure XP beads (Beckman Coulter Inc, Fullerton, CA, USA). Libraries were then normalized on specific beads according to the Nextera XT protocol (Illumina Inc, San Diego, CA, USA) and pooled. Automated cluster generation and pairwise sequencing with dual-index reads were performed in 2×250 bp with the Miseq Reagent Kit (V2-500 cycles) (Illumina Inc, San Diego, CA, USA). We chose to sequence wastewater samples on a MiSeq Illumina instrument to avoid possible cross contaminations with all clinical samples received in our laboratory and routinely sequenced on the NovaSeq6000 instrument.

Viral genomes from clinical samples were sequenced on a NovaSeq 6000 instrument (Illumina Inc., San Diego, CA, USA), as previously described [26].

### Sequences analysis

Reads of wastewater samples were analyzed as previously described [27]. Briefly, reads from pool1 and pool2 provided by the ARTIC procedure were mapped together against the Wuhan-Hu-1 SARS-CoV-2 isolate genome (GenBank accession number NC_045512.2) using the CLC genomics softwarev7.5 (Qiagen Digital Insights, Germany) with the default parameters. The non-synonymous mutations present in more than 10% of the reads were taken into account. For each sample, non-synonymous mutations were individually compared with classifying mutations that matched with 225 SARS-CoV-2 variants and sub-variants that had circulated since the beginning of the pandemics, including those that were circulating at this time, we call here the non-synonymous mutations mapped during the analysis: mutations patterns. When a mutation patterns occurred in more than one variant or sub-variant, all variants and sub-variants were added to the results (Supplementary Table 1).

For clinical samples, <u>g</u>enome consensus sequences were generated with the CLC Genomics workbench v.7 by mapping on the SARS-CoV-2 genome GenBank accession no.NC_045512.2 with the following thresholds: 0.8 for coverage and 0.9 for similarity. Sequences from complete genomes and clade assignment were analyzed using the Nextclade web-tool (https://clades.nextstrain.org/) [28].

## RESULTS

Wastewater 2212 was screened positive with cycle threshold value (Ct) of 31.2 and 34.4 for systems targeting the E gene and the mutation E484A respectively. Wastewater 2412 was also positive with Ct of 32.7 and 34.9 for systems targeting the E gene and the mutation E484A respectively.

For the sample 2212, on the 1,419,298 reads obtained, 97.3% were mapped, covering 75% of the reference genome. When considering a threshold of 10%, 31 non-synonymous mutations were present, 14 of them being signature mutations of SARS-CoV-2 variants (Figure 1). Besides, P4715L and D614G present in the majority of SARS-CoV-2 variants were found in 100% of the reads. Of 27 cumulative Omicron BA.1/21K and BA.1.1 subvariant mutations covered by the reads, 13 were present at a frequency ranging from 51 to 100%. The following specific mutations of these subvariants K856R, S3673_G3676S, I3758V, T547K, N856K were found. Of 20 and 32 mutation patterns of the Omicron BA.1.1.529 and BA.2/21L subvariants covered by the reads, 8 were found at a frequency ranging from 17 to 100%.

**Figure 1.**
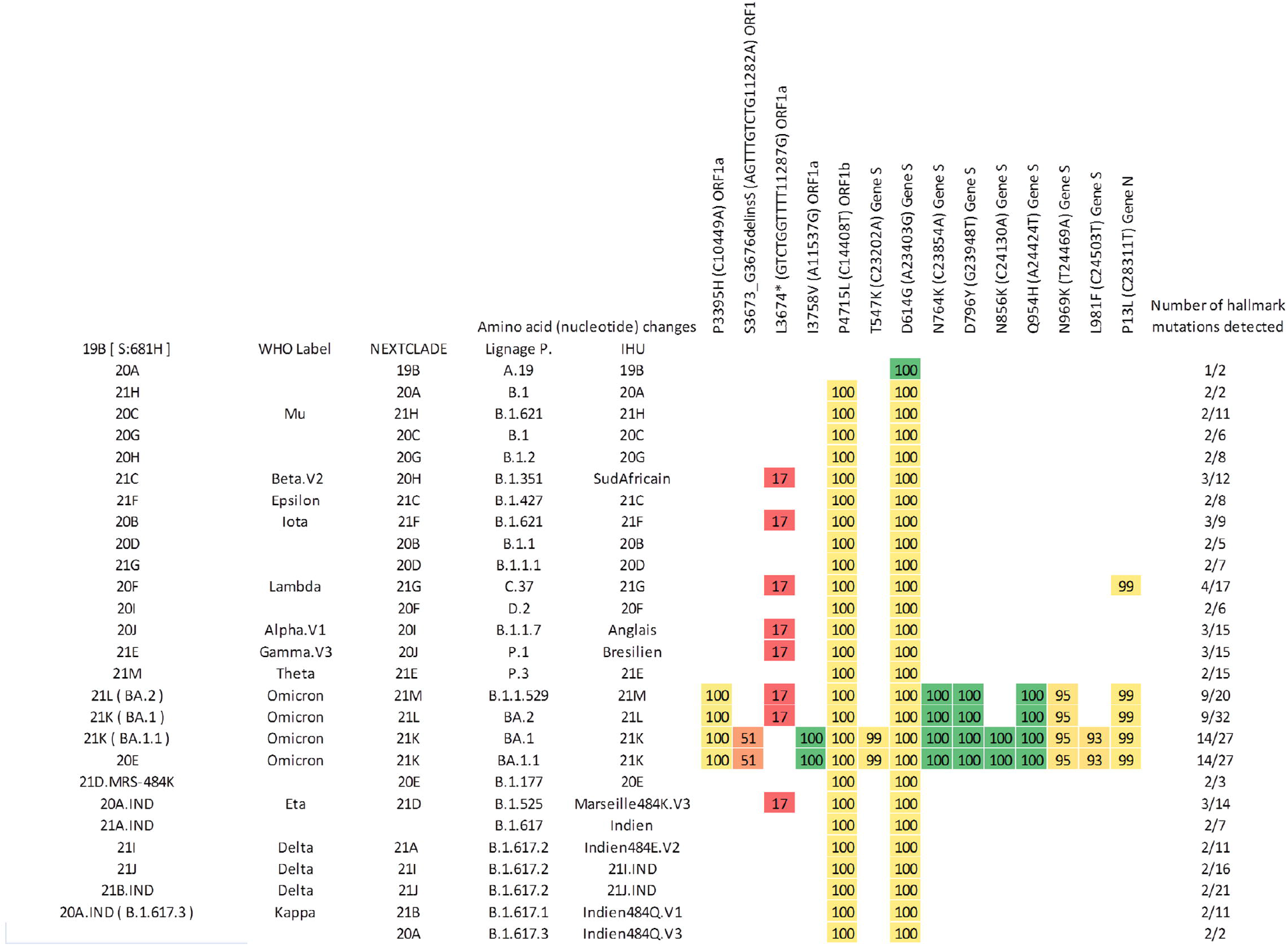
Mutation matrix, showing the mutations of the samples and the variants for which these mutations are found. The number indicated in the orange cells is the frequency of detection of the mutation. These results were obtained from the mapping of the sample 2212 to the SARS-CoV-2 Wuhan-Hu-1 isolate genome (GenBank accession number NC_045512.2) using the CLC genomics software v7.5 (Qiagen Digital Insights, Germany) with default settings. Only non-synonymous mutations present on at least 10% of reads were recovered. Variant labels include our local nomenclature (implemented at IHU Méditerranée Infection), and Nextstrain and Pangolin nomenclatures.

A total of 1,274,982 reads were obtained for the sample 2412, of which 83.7% were mapped, covering 71.6% of the reference genome. Twenty-four non-synonymous mutations were present when considering a threshold of 10%. The analysis of the reads revealed 13 non-synonymous mutation patterns specific of variants (Figure 2). D614G mutation was found in 100% of the reads. Of the 34 Omicron BA.1 and BA.1.1 mutation patterns covered by the reads, 9 were present with a frequency of 36 to 100%. The K856R, S2083_L2084delinsIle, S3673_G3676S andT547K mutations specific of these sub-variants were found. On a total of 27 and 39 mutation patterns of Omicron BA.1.1.529 and BA.2 sub-variants covered by the reads, 6 were present with a frequency ranging from 25 to 100%. Six mutation patterns specific of the Delta variant (comprising between 10 and 18 mapped mutations) were present with a frequency ranging from 11 to 100%.

**Figure 2.**
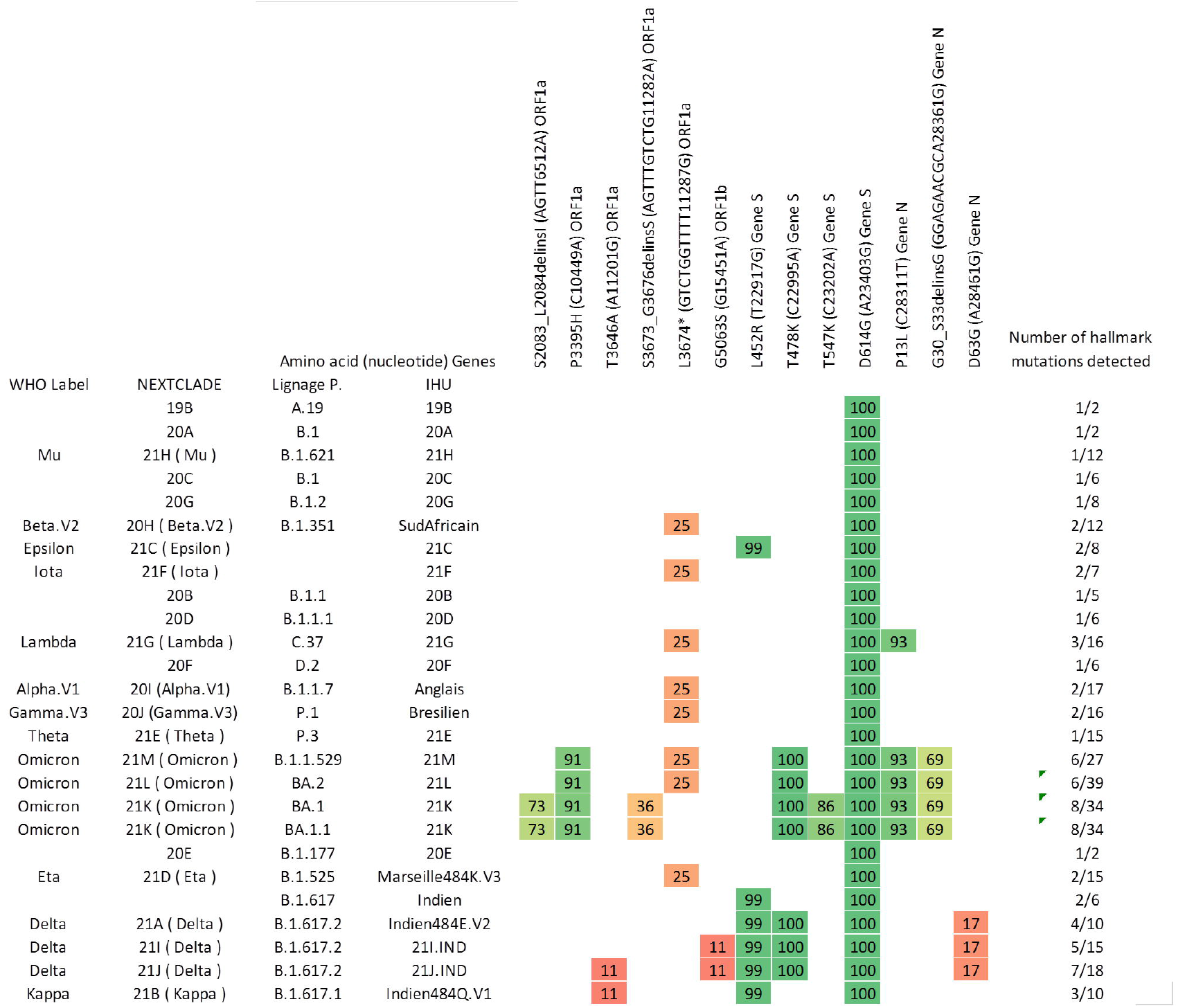
Mutation matrix, showing the mutations of the samples and the variants for which these mutations are found. The number indicated in the orange cases is the occurrence frequency of the detection of the mutation. These results were obtained from the mapping of the sample 2412 to the reference SARS-CoV-2 genome. (GenBank accession number NC_045512.2) using the CLC genomics software v7.5 (Qiagen Digital Insights, Germany) with default settings. Only non-synonymous mutations present on at least 10% of reads were recovered. Variant labels include the local nomenclature (IHU) and Nextstrain, and Pangolin nomenclatures.

For the aircraft that flew on December 24th, 2021, among the 56 passengers tested at the exit of the plane, by a rapid antigenic diagnostic test, 12 (21%) were detected as positive. The result was obtained in 20 minutes and directly communicated to the passengers. Among them, 11 were confirmed byRT-PCR. SARS-CoV-2 next-generation genome sequencing with the COVIDSeq protocol performed for these 11 samples provided the identification of the Omicron BA.1 variant for all.

## DISCUSSION

In this work, we screened the aircraft wastewater of the flight from Ethiopia to Marseille. RT-PCR screening showed the presence of the Omicron variant. The combined ARTIC and Illumina sequencing revealed the presence of mutation patterns of the Omicron variant. Interestingly, 13 passengers from the flight of December 24th, 2021 were tested positive by rapid antigenic test and the full genome sequencing of 11 of them revealed the presence of the Omicron variant. Thus, despite a negative PCR test requirement performed in the previous 72 hours before boarding as required even for vaccinated French people, one quarter of all passengers on board were infected. Besides, the congruent results for qPCR screening and variant detection by NGS on aircraft wastewater and the genotype obtained on clinical samples showed that aircraft wastewater surveillance by NGS is efficient for monitoring the circulation of variants, and especially, a possible powerful strategy for preventing massive importation of new variants of concern from abroad.

Monitoring of SARS-CoV-2 circulation in wastewater has already proven to be an effective tool for tracking infections at the community level, correlated with the number of individual cases [29]. Such approach applied to the aircraft wastewater may be a powerful tool for controlling SARS-CoV-2 importation and exportation, this risk existing despite strict control measures of passengers by mandatory clinical negative testing. Recent studies have shown that SARS-CoV-2 monitoring in wastewater from international flights and cruise ships is useful to prioritize testing of onboard passengers, and to improve management of contact tracing [20;22]. Ahmed et al recently detected the Omicron variant by NGS of aircraft wastewater samples collected from flight arriving to Darwin (Australia) from Johannesburg, South Africa) [23]. They combined the ARTIC approach with the Oxford Nanopore-GridION technology and the ATOPlex combined with the DNBseq-g400 sequencing. In the present study, we also detected the Omicron variant in wastewater samples from a long-haul flight from Addis-Ababa to Marseille that lasted 9 hours. One limit of this study is the possible contamination by remaining traces of SARS-CoV-2 RNA from previous flights in blackwater tanks. However, results obtained here show that this risk is very limited given the good correlation with clinical testing results. Also, SARS-CoV-2 infection was not assessed for the aircraft crew.

The Omicron variant was designated on November 26^th^, 2021 as a variant of concern by the world health organization, and described as highly transmissible, with a potential of immune escape as assessed by a reduced efficiency of the protective immunity developed after COVID-19 vaccination. Within three weeks after the first declared cases in Botswana, it had been detected in 87 countries [30]. Similarly, a previous work carried out in our laboratory showed that of sixteen SARS-CoV-2 variants identified in Marseille since the beginning of pandemics, seven were imported through travel from abroad [1]. History as a lesson, to be early aware of and implement preventing measures for new potential future pandemic waves due to new emerging variants, tracking variants’ circulation is indispensable. The importation in Europe of the Omicron variant from South-Africa [30] perfectly illustrates that the airway traffic is a threatening and powerful entry point for new variants despite public health policies including the green passport and the RT-PCR test required for travelers (wrong certificates). This work shows that despite a requirement of a negative RT-PCR test result performed in the previous 72 hours before crossing the border, 20% of passengers were positive, highlighting a great failing in such prevention policy. Aircraft wastewater screening can be performed in one hour, a timeframe short enough to inform passengers before custom clearance, and for triggering nasopharyngeal testing and strict quarantine until results. Such approach could be an efficient tool, probably more powerful than the mandatory certificate of negative testing that can be falsified. Aircraft and cruise wastewater monitoring may be of particular interest during inter-epidemic periods and for remote territories, for which massive importation of new SARS-CoV2 variants would have considerable public health consequences. Based on these findings, we propose wastewater SARS-CoV2 screening followed by variants’ monitoring by NGS as a global strategy for preventing new SARS-CoV-2 variants importation in unaffected regions, especially in isolated territories as islands or during periods of low viral circulation. Systematic screening and NGS on aircraft, and boat wastewater may help policy makers for targeted management strategies by testing and isolating passengers in case of a positive detection of SARS-CoV-2 in wastewater.

## Data Availability

All data produced in the present study are available upon reasonable request to the authors.

## Author contributions

LLT analyzed the data and wrote the manuscript. NW wrote the manuscript. AL collected the samples and performed PCR screening on wastewater and clinical samples. GP prepared samples for NGS and wrote the manuscript. PJ performed NGS and wrote the manuscript. PC revised the manuscript. BLS conceived the project, revised the manuscript and supervised the work. SA conceived data analysis, wrote the manuscript and supervised the work.

## Funding

This research was funded by the French Government under the “Investissements d’avenir” (Investments for the Future) program managed by the Agence Nationale de la Recherche (ANR, French National Agency for Research), reference: Méditerranée Infection 10-IAHU-03.

## Conflict of interest statement

Lorlane Le Targa is employee of Biosellal society.

## Ethics

Since July 2021, according to decree No. 2020-551 of 12 May 2020 on the information systems mentioned in Article 11 of Law No. 2020-546 of 11 May 2020 extending the state of health emergency for people arriving from countries classified at risk if the virus circulates actively, all passengers arriving at Marseille airport are proposed to have a naso-pharyngeal swab for SARS-CoV-2 antigenic detection according to a joint initiative of the regional prefecture and the regional health agency. Sampling and antigenic detection were performed by the bataillon des marins pompiers de Marseille. The positive samples collected for diagnostic purposes were reused for retrospective genotyping anonymously. According to French law (loi Jardé), anonymous retrospective studies do not require institutional review board approval nor informed consent. This study was approved by the Ethics Committee of the IHU Mediterranée Infection under the number 2022–013.

## Supplementary files

**SF1**. Table of variants

**SF2**. Complete mutation matrix, showing all the signature mutations of the SARS-CoV2 variants, for which at least one signature mutation was detected in the wastewater 2212 sample. Grey cases are signature mutations of the SARS-CoV2 variant indicated in the matching line. Orange cases are signature mutations of the SARS-CoV2 variant indicated in the matching line and detected in the 2212 sample. The number indicated in the orange cases is the occurrence frequency of the detection of the mutation. These results were obtained from the mapping of the sample 2212 to the reference SARS-CoV-2 genome. (GenBank accession number NC_045512.2) using the CLC genomics software v7.5 (Qiagen Digital Insights, Germany) with default settings. Only non-synonymous mutations present on at least 10% of reads were recovered. Variant labels include the local nomenclature (IHU) and Nextstrain, and Pangolin nomenclatures.

**SF3**. Complete mutation matrix, showing all the signature mutations of the SARS-CoV2 variants, for which at least one signature mutation was detected in the wastewater 2412 sample. Grey cases are signature mutations of the SARS-CoV2 variant indicated in the matching line. Orange cases are signature mutations of the SARS-CoV2 variant indicated in the matching line and detected in the 2412 sample. The number indicated in the orange cases is the occurrence frequency of the detection of the mutation. These results were obtained from the mapping of the sample 2412 to the reference SARS-CoV-2 genome. (GenBank accession number NC_045512.2) using the CLC genomics software v7.5 (Qiagen Digital Insights, Germany) with default settings. Only non-synonymous mutations present on at least 10% of reads were recovered. Variant labels include the local nomenclature (IHU) and Nextstrain, and Pangolin nomenclatures.

## Notes

### Competing Interest Statement

Lorlane Le Targa is employee of Biosellal society.
Bernard La Scola is a scientific consultant for GIS Edem solution.

### Funding Statement

This research was funded by the French Government under the Investments for the Future program managed by the Agence Nationale de la Recherche (ANR, French National Agency for Research), reference: Mediterranee Infection 10 IAHU 03.

### Author Declarations

Since July 2021, according to decree No. 2020 551 of 12 May 2020 on the information systems mentioned in Article 11 of Law No. 2020 546 of 11 May 2020 extending the state of health emergency for people arriving from countries classified at risk if the virus circulates actively, all passengers arriving at Marseille airport are proposed to have a naso-pharyngeal swab for SARS CoV 2 antigenic detection according to a joint initiative of the regional prefecture and the regional health agency. Sampling and antigenic detection were performed by the bataillon des marins pompiers de Marseille. The positive samples collected for diagnostic purposes were reused for retrospective genotyping anonymously. According to French law (loi Jarde), anonymous retrospective studies do not require institutional review board approval nor informed consent. This study was approved by the Ethics Committee of the IHU Mediterranee Infection under the number 2022 013.

